# Coronavirus Disease (COVID-19) Global Prediction Using Hybrid Artificial Intelligence Method of ANN Trained with Grey Wolf Optimizer

**DOI:** 10.1101/2020.10.22.20217604

**Authors:** Sina Ardabili, Amir Mosavi, Shahab S. Band, Annamaria R. Varkonyi-Koczy

## Abstract

An accurate outbreak prediction of COVID-19 can successfully help to get insight into the spread and consequences of infectious diseases. Recently, machine learning (ML) based prediction models have been successfully employed for the prediction of the disease outbreak. The present study aimed to engage an artificial neural network-integrated by grey wolf optimizer for COVID-19 outbreak predictions by employing the Global dataset. Training and testing processes have been performed by time-series data related to January 22 to September 15, 2020 and validation has been performed by time-series data related to September 16 to October 15, 2020. Results have been evaluated by employing mean absolute percentage error (MAPE) and correlation coefficient (r) values. ANN-GWO provided a MAPE of 6.23, 13.15 and 11.4% for training, testing and validating phases, respectively. According to the results, the developed model could successfully cope with the prediction task.

## I. Introduction

COVID-19 disease broke out in December 2019 in Wuhan, China [1]. COVID-19 can be considered as a new virus that has no vaccination and proper medicine for treatment. Proper prediction strategy for the COVID-19 outbreak can attract attention to the strategies of quarantine and other governmental measures, like lockdown, media coverage on social isolation, and improving the public hygiene to control it [2]. Recently, several strategies, including mathematical models, have been successfully employed for pandemic prediction of disease. Singhal et al. (2020) used the Fourier decomposition method (FDM) and susceptible-infected-recovered (SIR) model for the prediction of the COVID-19 pandemic. According to the results, the total number of cases and death was predicted to be 12.7 × 106 and 5.27 × 105, respectively [3]. Sarkar et al. (2020) employed a mathematical model for the pandemic prediction of the COVID-19 in India. The proposed model was a combination of susceptible (S), recovered (R), asymptomatic (A), infected (I), quarantined susceptible (Sq), and isolated infected (Iq) models [4]. Anirudh (2020) employed SIR, SEIRU, SEIR, SLIAR, SIRD, ARIMA and SIDARTHE for the COVID-19 pandemic prediction [5]. But, mathematical models have disadvantages such as complexity, time-consuming and lower reliability [6, 7].

Machine Learning (ML) based models have been successfully employed in different fields of science [8]. Pandemic prediction can be considered one of the ML-based models’ fields that provided good accuracy and reliability. Recently, researchers have developed studies for the prediction of COVID-19 cases and death rate using ML-based methods. Tabrizchi et al. (2020) employed a Deep Learning-based estimation model integrated by an image-based diagnosis method to detect coronavirus infection [9]. Dutta et al. (2020) used a DL based prediction model for the estimation of the COVID-19 outbreak [10]. Ardabili et al. (2020) employed different single and hybrid ML-based methods to predict the COVID-19 epidemic in USA, Germany, China, Iran and Italy. According to the results, ML-based prediction models could successfully cope with the prediction task [11]. Pinter et al. (2020) employed hybrid ML-based models (ANFIS and ANN-ICA) for the prediction of total cases of COVID-19 in Hungary using time-series data [6]. Accordingly, the present study aimed to predict the global COVID-19 outbreak using a robust hybrid Artificial Neural Network integrated by Grey Wolf Optimizer (ANN-GWO) in the presence of the time-series total and daily COVID-19 cases.

## II. Methodology

The required data for developing the prediction model was generated from https://www.worldometers.info/. The nature of the dataset is time-series based. Accordingly, the total and the daily number of cases are presented in Figure I from January 22 to November 15, 2020. Figure I has two vertical axes. One is for total cases and the other is related to the daily cases.

Due to the use of time-series data, it has been decided to choose the input dataset according to Table I for the prediction of the global COVID-19 outbreak as the output parameter.

**TABLE I.**
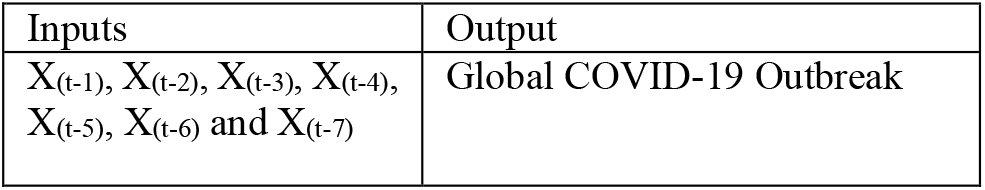
description of the inputs and output of modeling process

ANN-GWO performed modeling in the presence of input and output datasets. ANN-GWO can be considered as one of the robust hybrid ML-based methods [12]. The modeling phase was started by employing the different architectures of ANN and different population sizes for GWO. Finally, according to the model’s accuracy, ANN with the architecture of 7-10-4-1 in the presence of population size 150 for GWO was selected as the best prediction model for the global COVID-19 prediction with a high accuracy. 70% of total data were chosen for the training phase, 30% of total data were employed for the testing phase and the predicted COVID-19 outbreak for September 16 to October 20, 2020 were selected for the validation phase. The description of ANN-GWO has been comprehensively presented in our previous study in [11]. Figure II presents the main schematic of the developed model.

According to Figure II, the main mechanism of the GWO is to update the bias and weights of the desired ANN architecture to reach the highest accuracy.

The performance and accuracy of the developed method was evaluated using the Mean Absolute Percentage Error (MAPE) and correlation coefficient (r) according to Equations 1 and 2:

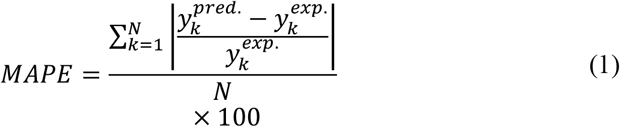

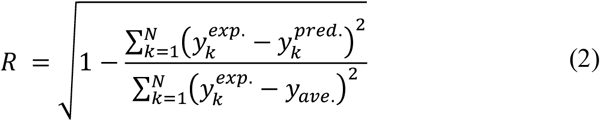

Where 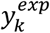 Refers to the target values, 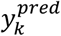 Refers to the predicted values and N refers to the number of data.

## III. Results

Table II presents the training results, testing and validating phases into two terms: MAPE and r values. According to the results, the selected ANN architecture with a 150 at the maximum iteration of 500 provided a good accuracy for training, testing and validating phases.

**TABLE II.** results of the training, testing and validating phases

| ANN architecture | Pop. size | Maximum iteration | Phase | MAPE | Correlation coefficient |
| --- | --- | --- | --- | --- | --- |
| 7-10-4-1 | 150 | 500 | Training | 6.23 % | 0.999 |
|  |  |  | Testing | 13.15% | 0.994 |
|  |  |  | Validating | 11.4% | 0.994 |

Figure III (a) and (b) present the plot diagram of the predicted and target values and the deviation from the testing phase’s target values. As is clear from Figure 3a and according to the testing phase’s determination coefficient, the testing phase’s performance is acceptable.

**Figure I.**
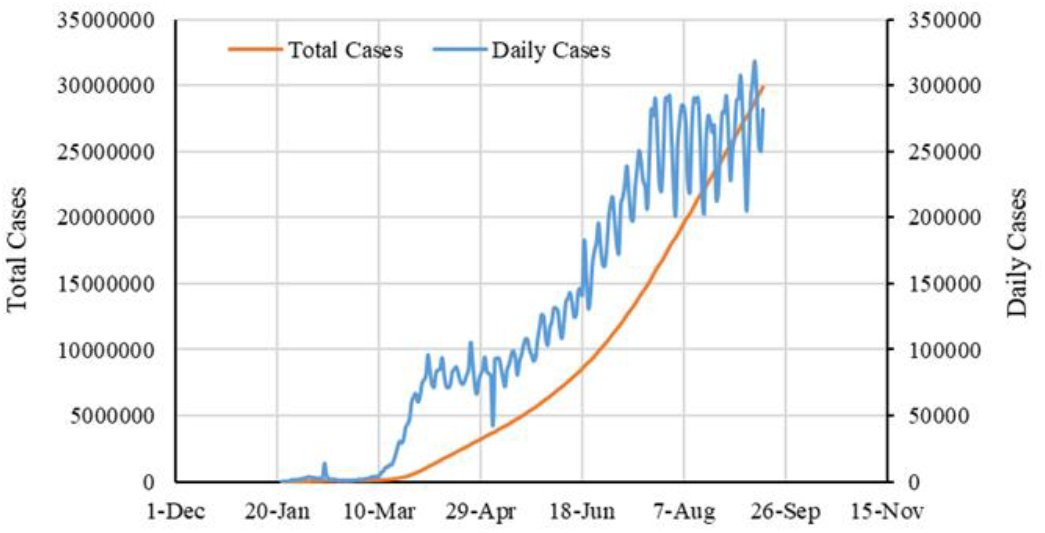
The employed time-series dataset for developing the ML-based prediction model

**Figure II.**
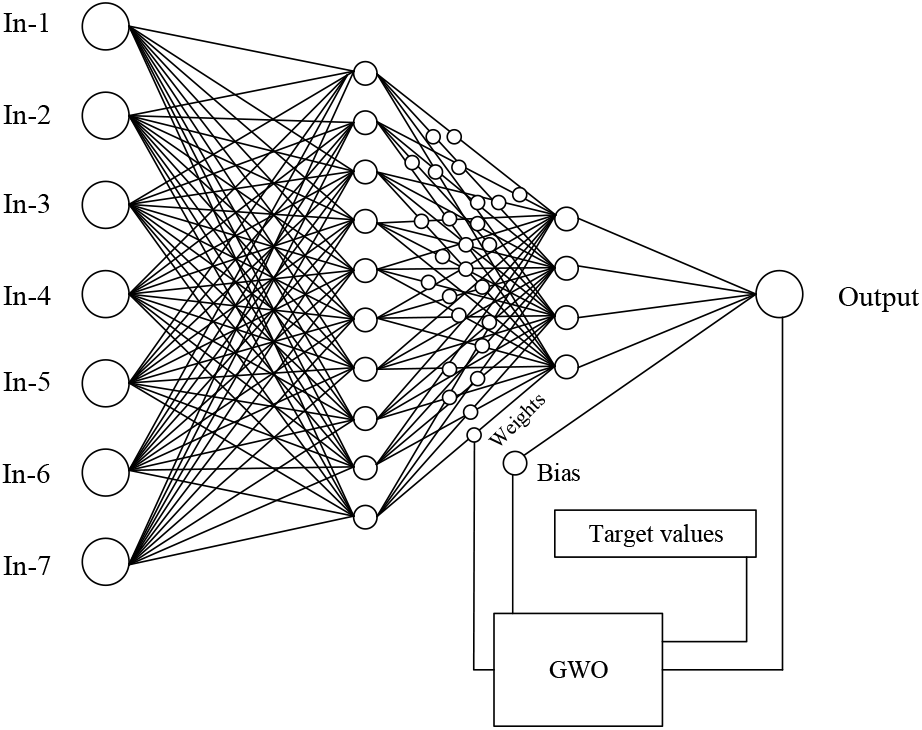
the main schematic of ann-gwo

**Figure III.**
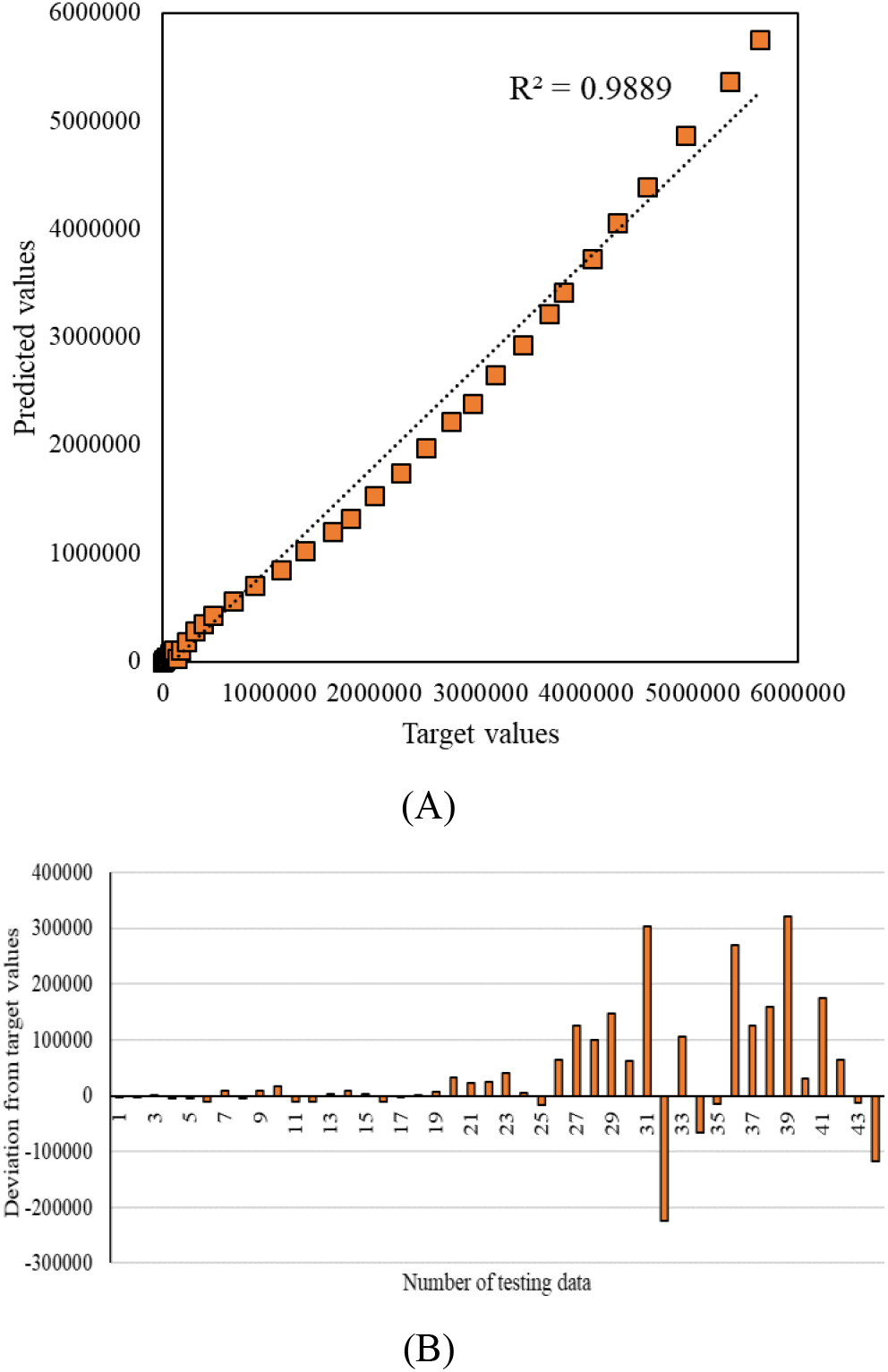
(A) THE PLOT DIAGRAM, (B) DEVIATION FROM TARGET VALUES.

Figure 4 presents the outbreak prediction by ANN-GWO from October 16 to June 4, 2021 (for 232 days. As is clear according to the projections, the outbreak of the COVID-19 is still increasing and the trend is rising. This can be a severe warning to the World Health Organization.

**Figure IV.**
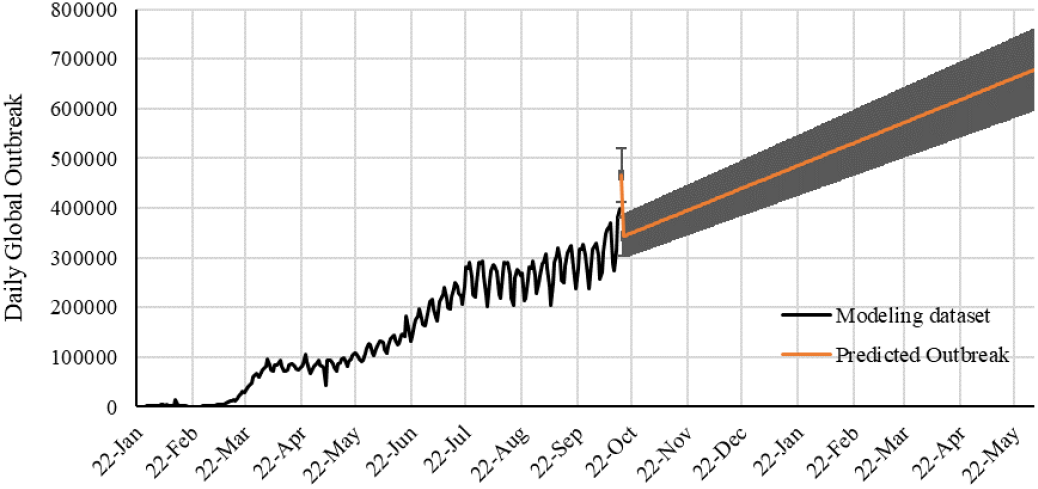
The prediction of the Global COVID-19 outbreak from October 16 to June 4, 2021

## IV. Conclusion

In this paper, the COVID-19 outbreak is modeled as a complex time series. We have developed a hybrid machine learning model based on the artificial neural network. We trained the system with the grey wolf optimization algorithm to get the highest performance. Based on the testing, we projected the outbreak until late May. Training and testing processes have been performed by time-series data related to January 22 to September 15, 2020 and validation has been performed by time-series data associated with September 16 to October 15, 2020. Results have been evaluated by employing mean absolute percentage error (MAPE) and correlation coefficient (r) values. ANN-GWO provided a MAPE of 6.23, 13.15 and 11.4% for training, testing and validating phases, respectively. According to the results, the developed model could successfully cope with the prediction task. The model provided promising results, and it can be used for outbreak prediction.

## Data Availability

https://www.worldometers.info/

## Acknowledgment

This research in part, is supported by the Hungarian State and the European Union under the EFOP-3.6.2-16-2017-00016 project. The support of the Alexander von Humboldt Foundation is acknowledged.

